# Inconsistent Music-Based Intervention Reporting in Dementia Studies: A Systematic Mapping Review

**DOI:** 10.1101/2024.02.21.24303087

**Authors:** Rebecca J. Lepping, Benjamin J. Hess, Jasmine M. Taylor, Deanna Hanson-Abromeit, Kristine N. Williams

## Abstract

Recent research has shown beneficial results for music-based interventions (MBIs) for persons living with Alzheimer’s disease and related dementias (AD/ADRD), but reports often lack sufficient detail about the MBI methodology, which reduces replicability. A detailed checklist for best practices in how to report MBIs was created in 2011 by Robb and colleagues to remedy the lack of detail in MBI descriptions. The implementation of the checklist specifically in AD/ADRD research has not been established. Given the complexity of music and the variety of uses for research and health, specific MBI descriptions are necessary for rigorous replication and validation of study results.

This systematic mapping review utilized the “Checklist for Reporting Music-Based Interventions” to evaluate the current state of MBI descriptive specificity in AD/ADRD research. Research articles testing MBIs and reviews of MBI efficacy published between January 2015 and August 2023 were scored using the checklist and the results were summarized. Nineteen studies were screened, and reporting was inconsistent across the 11 checklist criteria. Six out of 19 studies fully reported more than 5 of the 11 criteria. Only one of the 11 scoring criteria was at least partially reported across all 19 studies.

Thorough reporting of intervention detail for MBIs remains limited in AD/ADRD MBI research. This impedes study validation, replication, and slows the progress of research and potential application of music in practice. Greater implementation of the reporting guidelines provided by Robb and colleagues would move the field of MBI research for AD/ADRD forward more quickly and efficiently.

## Inconsistent Music-Based Intervention Reporting in Dementia Studies

Music-based interventions for health and wellbeing are receiving increased attention due in part to their lower cost, broader accessibility, and minimal side effects relative to pharmacological interventions. There is favorable evidence from recent research that MBIs are beneficial for people with AD/ADRD. One hypothesized reason for the effectiveness of MBIs is the observation that musical memory is retained and can still evoke a response throughout the progression of the disease, even in later stages when communication becomes more difficult [1–4]. Multiple review papers have concluded that MBIs produce beneficial outcomes for people with AD/ADRD [2–15], while others have conceded that beneficial outcomes are probable although the direct evidence may be weak [16–18]. Some benefits MBIs provide for people with AD/ADRD are reduced stress, reduced emotional disturbances and depression, and improved memory and cognitive function [1, 2, 4, 6, 8, 10, 13, 14, 16, 18, 19]. These benefits may also extend to caregivers of people with AD/ADRD, who also often experience a decrease in quality of life as they care for their loved ones [8, 10, 20].

The interest in the effectiveness of MBIs as a non-pharmacological treatment continues to grow, and evidence for their beneficial effects is favorable. However, specific features of the music need to be consistently identified and described to move music-based interventions from anecdotal evidence into the realm of prescriptive interventions. A wide variety of funding opportunities which allow for the incorporation of music-based interventions, including one specifically focused on funding MBI research, are available and will likely continue to fuel this increase in MBI research [21]. However, the mechanisms which produce the beneficial effects of MBIs are not always clearly defined or understood. Understanding the underlying biological or psychological mechanism likely to be affected by the MBI gives a clearer understanding of which aspect of the intervention is producing the effect. This is also an area of interest for the NIH. Funding for dementia studies with a specific focus on the underlying mechanisms is also available, a fact which further supports both the level of importance placed on clearly defined mechanistic understanding and the necessity for more detailed and rigorous research [22].

The importance of choosing specific music elements to focus on when designing and describing an intervention has been similarly highlighted by the NIH and others. Musical elements and the ways in which humans respond to these elements are both complex. This complexity requires clear descriptions of the hypothesized interactions when designing an intervention and clear reporting of the musical elements used and the methods which drive their selection. Clear description of these elements is necessary, not only to define the mechanism of the intervention and interpret results; but also to aid in reproducing the effect in future studies. Establishing guidelines and frameworks for reporting is an essential part of achieving clear reporting, which has been recently provided by the NIH in the Music-Based Intervention Toolkit and in the Therapeutic Function of Music framework outlined by Hanson-Abromeit [23, 24].

Previously published reviews have focused on the results of music-based interventions for AD/ADRD, but few have considered the potential variability of the music interventions themselves resulting from the lack of detailed and specific descriptions. Differences and similarities between MBIs from one study to the next are difficult to determine because they are often only vaguely described. Without specific descriptions of the qualities of the music elements within music stimuli, the MBIs cannot be accurately reproduced, limiting the conclusions that can be drawn regarding efficacy. Unfortunately, reproducing individual study results in subsequent trials has proven difficult given the inconsistent levels of detail used to describe the music interventions across studies.

This gap was identified in 2011 by Dr. Sheri Robb and colleagues, who described the need for consistent and specific MBI reporting standards across inter-disciplinary research on music-therapy interventions [25]. This team of music therapists and researchers created the “Checklist for Reporting Music-Based Interventions” to assist future researchers and improve transparency and rigor in music-based intervention research [25]. In a 2018 follow-up review, Robb and colleagues examined the reporting specificity, based on their checklist, of MBI studies from 2010-2015 across a wide range of disciplines in healthcare. The result of their study was that consistent detailed reporting was not observed [26]. To map the quality of reporting for MBIs specifically in AD/ADRD research since the previous review, we conducted an updated systematic mapping review of reporting rigor for MBI research studies in AD/ADRD from 2015-2023. For the purposes of this review, we searched for studies published since 2015 to identify studies published after the previous 2018 review by Robb and colleagues. Our aim was to discover whether reporting of MBIs for people with AD/ADRD had improved since the evaluation conducted up to 2015, and to describe any consistencies and inconsistencies that we observed in recent reporting.

## MATERIALS AND METHODS

A systematic mapping review was conducted, searching PubMed on August 22, 2023 for papers published between 2015-2023 related to music interventions and Alzheimer’s or dementia. The search terms used, inclusion, and exclusion criteria are in Table 1. The results of these searches were filtered, using the PubMed search filters, to include only those published between 2015-2023, with free full text in English available that were either meta-analysis, review, systematic review, or randomized controlled trials. The resulting paper titles were screened for inclusion based on our inclusion and exclusion criteria (Table 1). Articles with titles referencing Alzheimer’s/dementia/cognitive decline and a music-based intervention were selected, and duplicate articles discovered across multiple searches were removed. The remaining articles were divided into review articles (meta-analysis, review, systematic review) and study articles (randomized controlled trial, prospective study). The review articles were then further screened based on full text review and level of relevance to MBI’s for AD/ADRD, those with the highest level of relevance were retained. Review results were used to evaluate whether MBI’s had beneficial outcomes, and what specific outcomes had been observed, because they synthesized a wide range of data that had already been reviewed for study quality. Each study article was further screened based on full text review and further study articles were then selected using the reference lists of the selected study articles. Screening was carried out by coauthor, BH.

**Table 1.** Search Terms, Inclusion, and Exclusion Critera.

|  |
| --- |
| ***Search Terms*** |
| “AD” OR “Alzheimer’s” AND “music” |
| “AD” AND “music” AND “intervention” |
| “Alzheimer’s” AND “music” AND “intervention” |
| “music” AND “dementia” |
| “music” AND “cognitive decline” |
| ***Inclusion Criteria*** |
| Articles published between January 1<sup>st</sup>, 2015 and August 22<sup>nd</sup>, 2023 |
| AD/ADRD focused |
| Uses music-based intervention |
| Randomized controlled trial, prospective study, meta-analysis, or review |
| Study was not included in the 2018 review by Robb and colleagues |
| ***Exclusion Criteria*** |
| No music-based intervention |
| Not AD/ADRD focused |
| Study was previously included in 2018 review by Robb and colleagues |

The study articles were evaluated to determine the specificity of MBI descriptions and whether they met the standards of the “Checklist for Reporting Music-Based Interventions” (Supplementary Table 1). Each of the qualifying studies was scored based on the checklist which has 7 items. One of these items consists of 5 sub-categories which were treated as separate items for the purpose of this analysis for a total of 11 scored items. These scores were used to identify patterns of reporting across studies. Each checklist item could receive one of three possible scores, based on whether the item was found anywhere in the MBI description or within the article. The three scores were 0 (not observed/described), 0.5 (partially observed/described), or 1 (fully observed/described). A score of 0 was assigned if no description of a checklist item could be found within the full text of the report. A score of 0.5 was assigned if a description of a checklist item was found within the full text of the report, but all the item components were not described. A score of 1 was assigned if a description of a checklist item was found within the full text of the report and all item components were described. The scoring was carried out independently by two raters (Rater initials: BH and AZ). Interrater scoring disagreements were reviewed and reconciled by a third reviewer (RL). The scoring was summed across checklist items and studies and visualized in Microsoft Excel to generate charts and observe qualitative patterns in the data. A PRISMA checklist for this systematic mapping review is provided in Supplementary Table 2 [27].

## RESULTS

The PubMed search resulted in the selection of 64 articles which were divided between 16 studies and 48 reviews. After full text examination of the studies, 3 articles failed to meet inclusion criteria and were excluded as ineligible. The reference sections of the remaining 13 study articles were searched for additional relevant literature; 6 additional study articles were identified and included for a combined total of 19 studies. The 48 reviews were then further screened based on full text review and level of relevance to MBI’s for AD/ADRD, those with insufficient relevance were removed which resulted in 19 total reviews. This screening was carried out by BH (Figure 1, Table 2).

**Figure 1.**
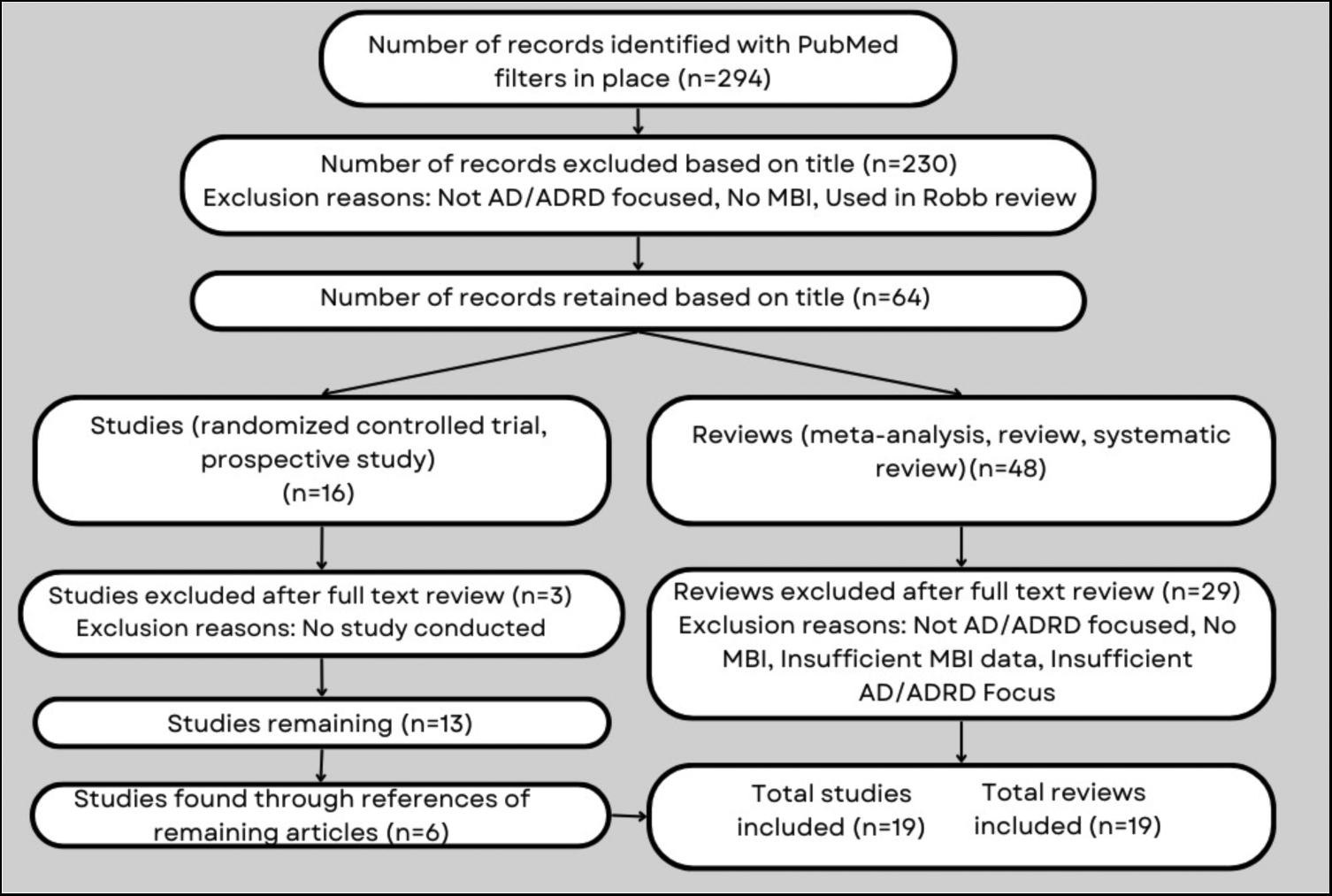
Search results flowchart.

**Table 2:**
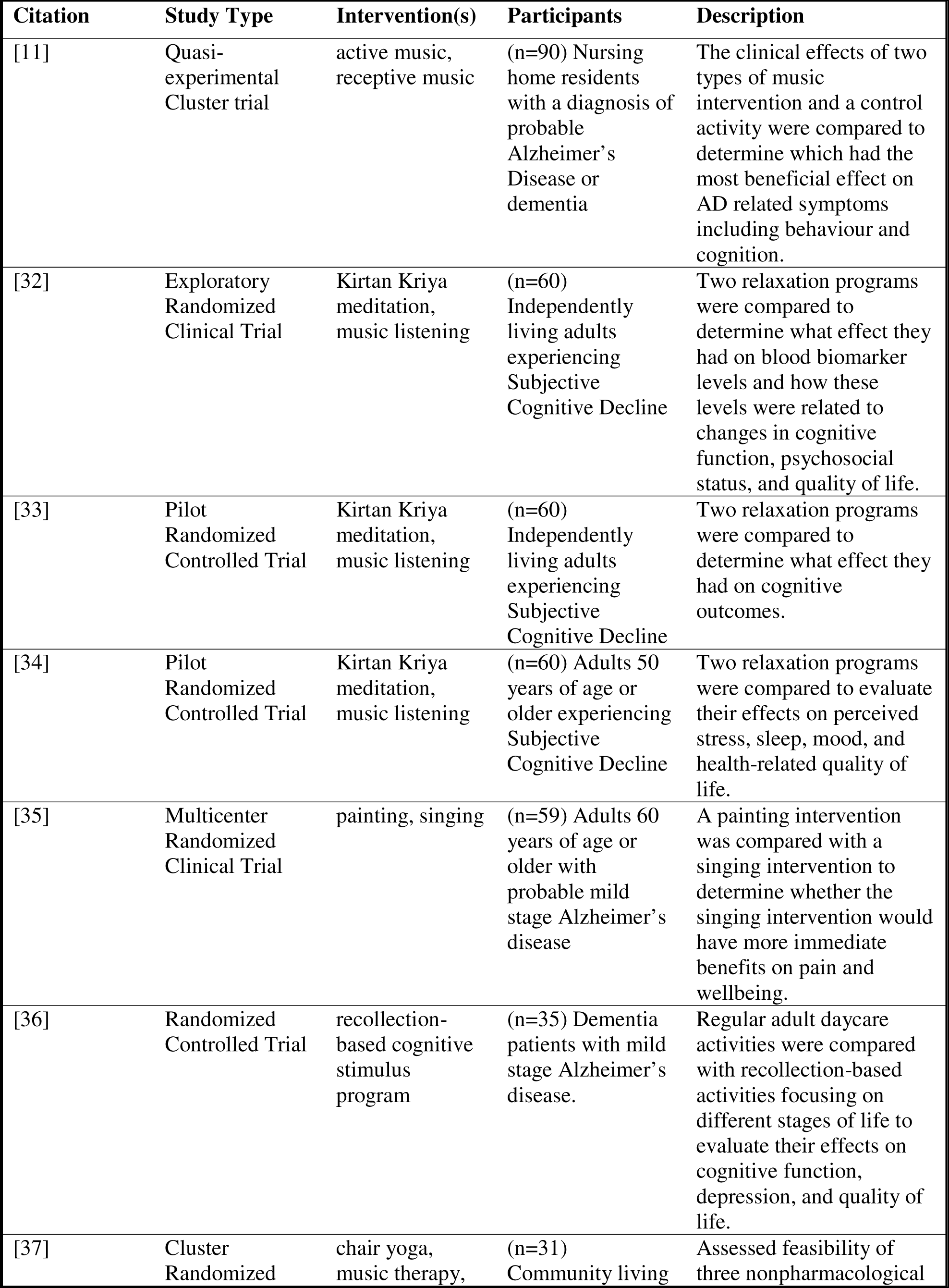
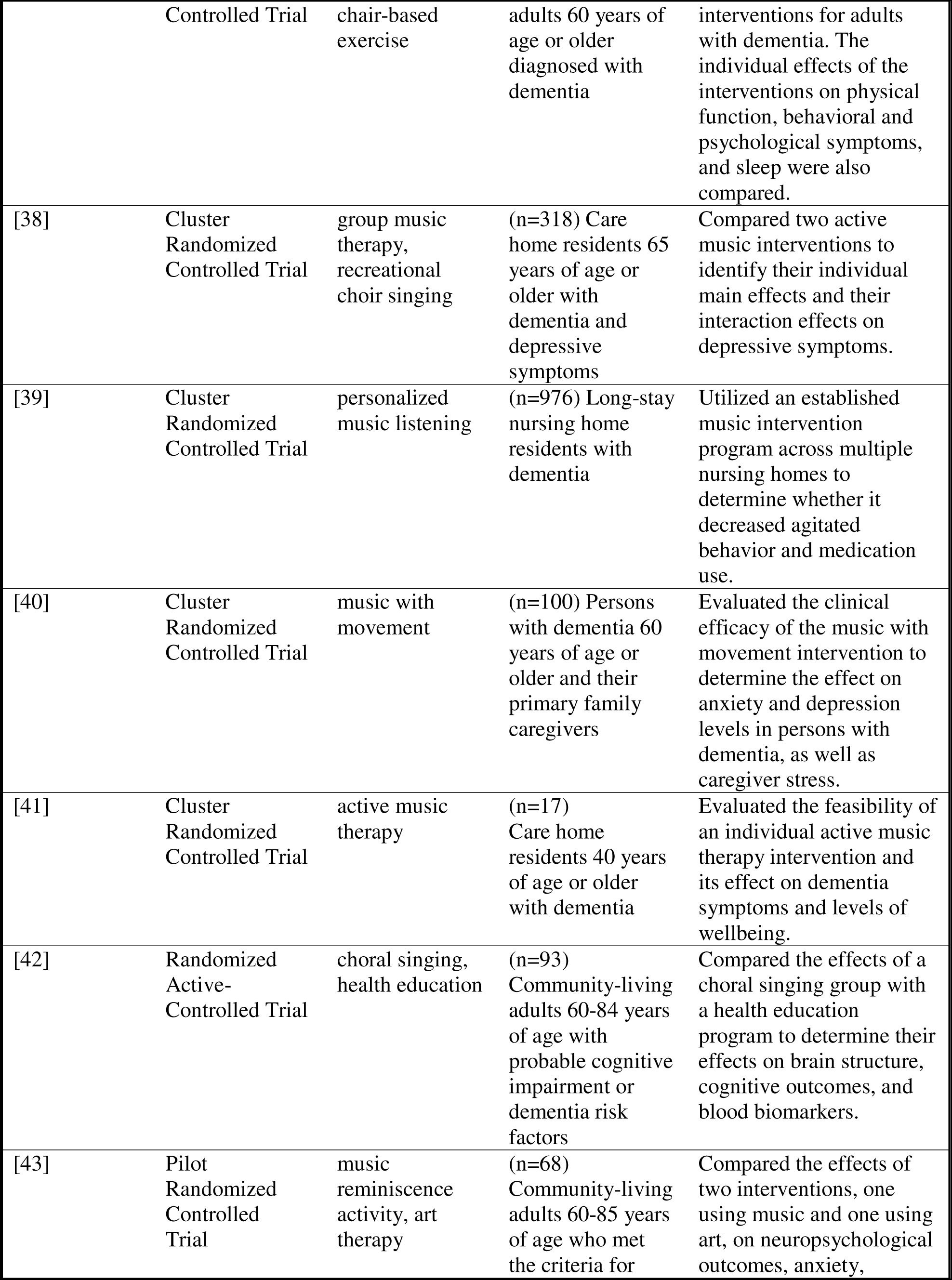
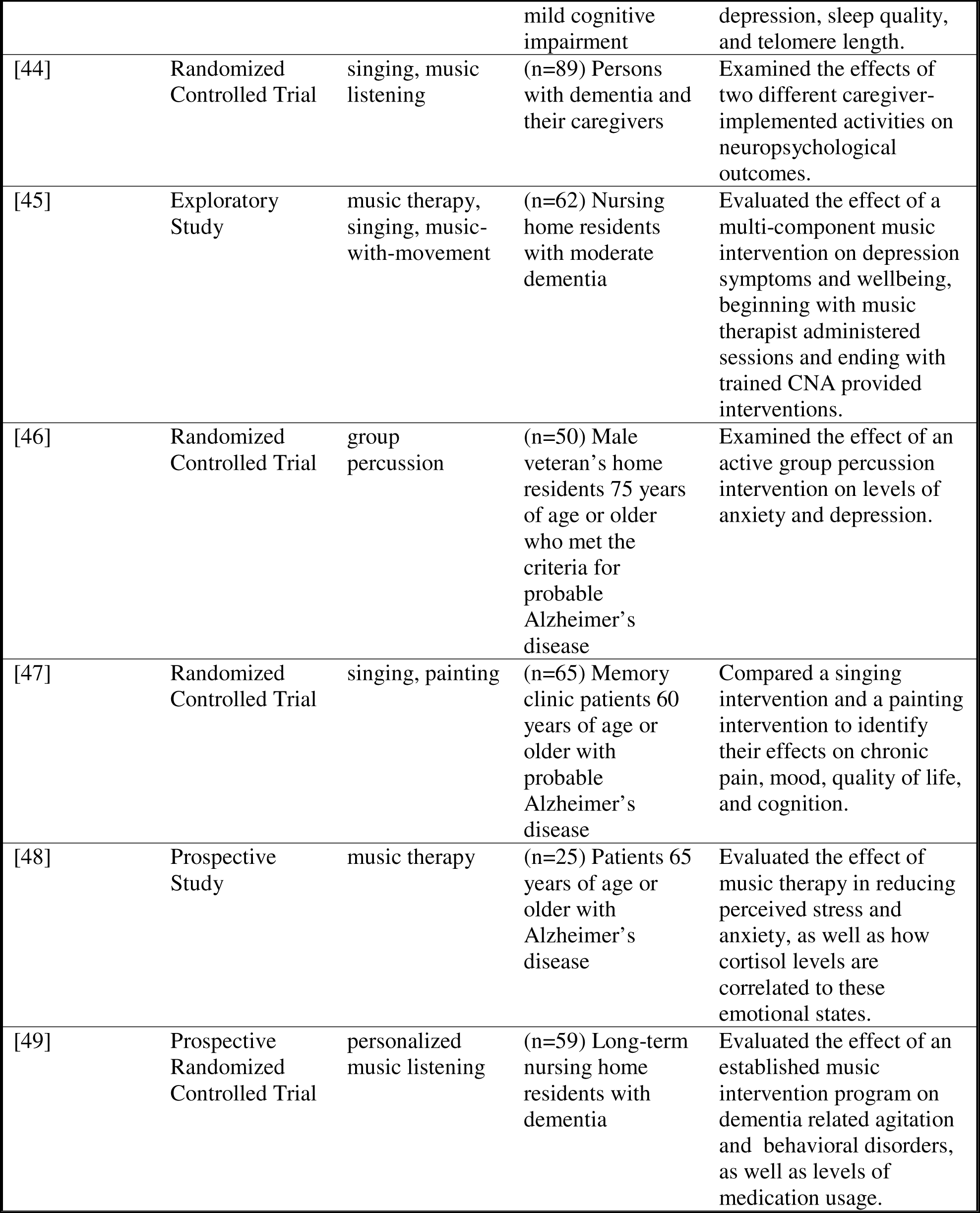
Studies evaluated for reporting specificity (n=19)

MUSIC INTERVENTION REPORTING IN DEMENTIA
| Citation | Study Type | Intervention(s) | Participants | Description |
| --- | --- | --- | --- | --- |
| [11] | Quasi-experimental Cluster trial | active music, receptive music | (n=90) Nursing home residents with a diagnosis of probable Alzheimer's Disease or dementia | The clinical effects of two types of music intervention and a control activity were compared to determine which had the most beneficial effect on AD related symptoms including behaviour and cognition. |
| [32] | Exploratory Randomized Clinical Trial | Kirtan Kriya meditation, music listening | (n=60) Independently living adults experiencing Subjective Cognitive Decline | Two relaxation programs were compared to determine what effect they had on blood biomarker levels and how these levels were related to changes in cognitive function, psychosocial status, and quality of life. |
| [33] | Pilot Randomized Controlled Trial | Kirtan Kriya meditation, music listening | (n=60) Independently living adults experiencing Subjective Cognitive Decline | Two relaxation programs were compared to determine what effect they had on cognitive outcomes. |
| [34] | Pilot Randomized Controlled Trial | Kirtan Kriya meditation, music listening | (n=60) Adults 50 years of age or older experiencing Subjective Cognitive Decline | Two relaxation programs were compared to evaluate their effects on perceived stress, sleep, mood, and health-related quality of life. |
| [35] | Multicenter Randomized Clinical Trial | painting, singing | (n=59) Adults 60 years of age or older with probable mild stage Alzheimer's disease | A painting intervention was compared with a singing intervention to determine whether the singing intervention would have more immediate benefits on pain and wellbeing. |
| [36] | Randomized Controlled Trial | recollection-based cognitive stimulus program | (n=35) Dementia patients with mild stage Alzheimer's disease. | Regular adult daycare activities were compared with recollection-based activities focusing on different stages of life to evaluate their effects on cognitive function, depression, and quality of life. |
| [37] | Cluster Randomized | chair yoga, music therapy, | (n=31) Community living | Assessed feasibility of three nonpharmacological |
|  | Controlled Trial | chair-based exercise | adults 60 years of age or older diagnosed with dementia | interventions for adults with dementia. The individual effects of the interventions on physical function, behavioral and psychological symptoms, and sleep were also compared. |
| [38] | Cluster Randomized Controlled Trial | group music therapy, recreational choir singing | (n=318) Care home residents 65 years of age or older with dementia and depressive symptoms | Compared two active music interventions to identify their individual main effects and their interaction effects on depressive symptoms. |
| [39] | Cluster Randomized Controlled Trial | personalized music listening | (n=976) Long-stay nursing home residents with dementia | Utilized an established music intervention program across multiple nursing homes to determine whether it decreased agitated behavior and medication use. |
| [40] | Cluster Randomized Controlled Trial | music with movement | (n=100) Persons with dementia 60 years of age or older and their primary family caregivers | Evaluated the clinical efficacy of the music with movement intervention to determine the effect on anxiety and depression levels in persons with dementia, as well as caregiver stress. |
| [41] | Cluster Randomized Controlled Trial | active music therapy | (n=17) Care home residents 40 years of age or older with dementia | Evaluated the feasibility of an individual active music therapy intervention and its effect on dementia symptoms and levels of wellbeing. |
| [42] | Randomized Active-Controlled Trial | choral singing, health education | (n=93) Community-living adults 60-84 years of age with probable cognitive impairment or dementia risk factors | Compared the effects of a choral singing group with a health education program to determine their effects on brain structure, cognitive outcomes, and blood biomarkers. |
| [43] | Pilot Randomized Controlled Trial | music reminiscence activity, art therapy | (n=68) Community-living adults 60-85 years of age who met the criteria for | Compared the effects of two interventions, one using music and one using art, on neuropsychological outcomes, anxiety, |

**Table 2: Studies evaluated for reporting specificity (n=19)**
|  |  |  |  |  |
| --- | --- | --- | --- | --- |
|  |  |  | mild cognitive impairment | depression, sleep quality, and telomere length. |
| [44] | Randomized Controlled Trial | singing, music listening | (n=89) Persons with dementia and their caregivers | Examined the effects of two different caregiver-implemented activities on neuropsychological outcomes. |
| [45] | Exploratory Study | music therapy, singing, music-with-movement | (n=62) Nursing home residents with moderate dementia | Evaluated the effect of a multi-component music intervention on depression symptoms and wellbeing, beginning with music therapist administered sessions and ending with trained CNA provided interventions. |
| [46] | Randomized Controlled Trial | group percussion | (n=50) Male veteran's home residents 75 years of age or older who met the criteria for probable Alzheimer's disease | Examined the effect of an active group percussion intervention on levels of anxiety and depression. |
| [47] | Randomized Controlled Trial | singing, painting | (n=65) Memory clinic patients 60 years of age or older with probable Alzheimer's disease | Compared a singing intervention and a painting intervention to identify their effects on chronic pain, mood, quality of life, and cognition. |
| [48] | Prospective Study | music therapy | (n=25) Patients 65 years of age or older with Alzheimer's disease | Evaluated the effect of music therapy in reducing perceived stress and anxiety, as well as how cortisol levels are correlated to these emotional states. |
| [49] | Prospective Randomized Controlled Trial | personalized music listening | (n=59) Long-term nursing home residents with dementia | Evaluated the effect of an established music intervention program on dementia related agitation and behavioral disorders, as well as levels of medication usage. |

It should be noted that none of the 19 studies included in this review cited the 2011 paper by Robb and colleagues which included the reporting checklist [25]. Consequently, the studies cannot be reasonably expected to include the checklist items verbatim. However, the checklist still serves as a valuable reference for determining how specific the descriptions were. The checklist items are “A: Intervention Theory”, “B: Intervention Content” (contains five sub-categories), “C: Intervention Delivery Schedule”, “D: Interventionist”, “E: Treatment Fidelity”, “F: Setting”, and “G: Unit of Delivery” [25]. The MBI descriptions in the study articles were limited, and full points were rarely awarded. One study received full points for eight items, two received full points for seven items, three received full points for six items, one received full points for five items, and the remaining 12 studies received full points for four or fewer items Figure 2 displays the total score each study received, with each bar section color coded to show the point value contributed by each checklist item. Full points and half points are indicated by the height of each column section. The maximum score each study could receive was 11. As seen in Figure 2, only six studies fully described more than five of the 11 items on the checklist, and when accounting for partial scoring, only nine of the 19 studies achieved a score exceeding 5.5 out of 11. Figure 3 shows the total score for each checklist item across all studies, each section of the mountain plot is color coded to identify which study contributed the point value, full points and half points are indicated by the height of the section. The maximum score each checklist item could receive was 19 (the number of studies). As shown in Figure 3, the most consistently described item was “C: Intervention Delivery Schedule”. Seventeen studies fully reported item C, and the remaining two partially reported item C. The least reported item was “B.2: Music”, with only one study partially reporting this item. Despite achieving a midrange score, item “A: Intervention Theory” was also underreported, with only seven studies fully reporting a theoretical rationale for how the MBI was hypothesized to effect change and eight out of 19 studies partially reporting this item.

**Figure 2.**
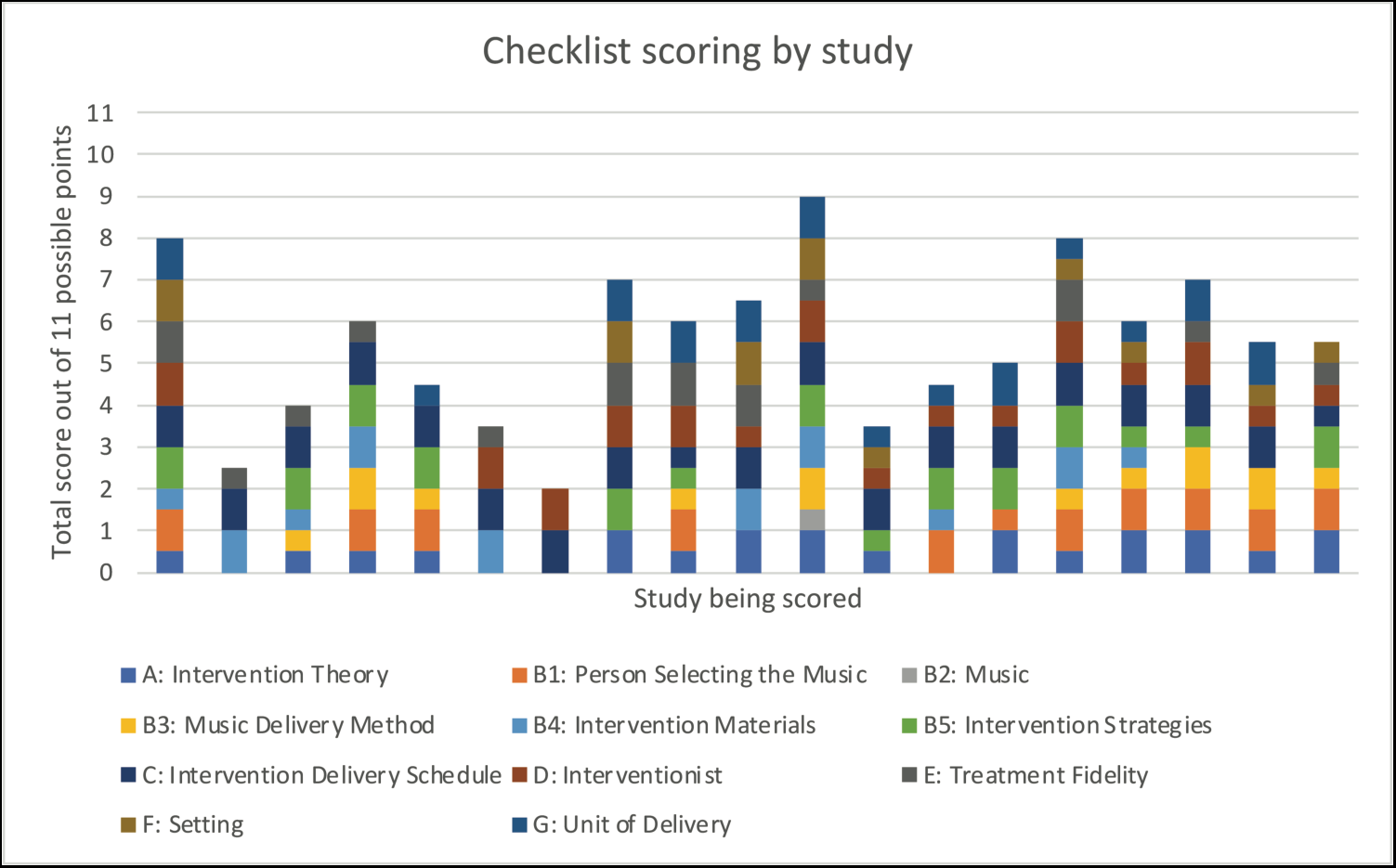
Number of checklist items reported in each study. Whole or half points are represented for each checklist item by the color coding described in the legend.

**Figure 3.**
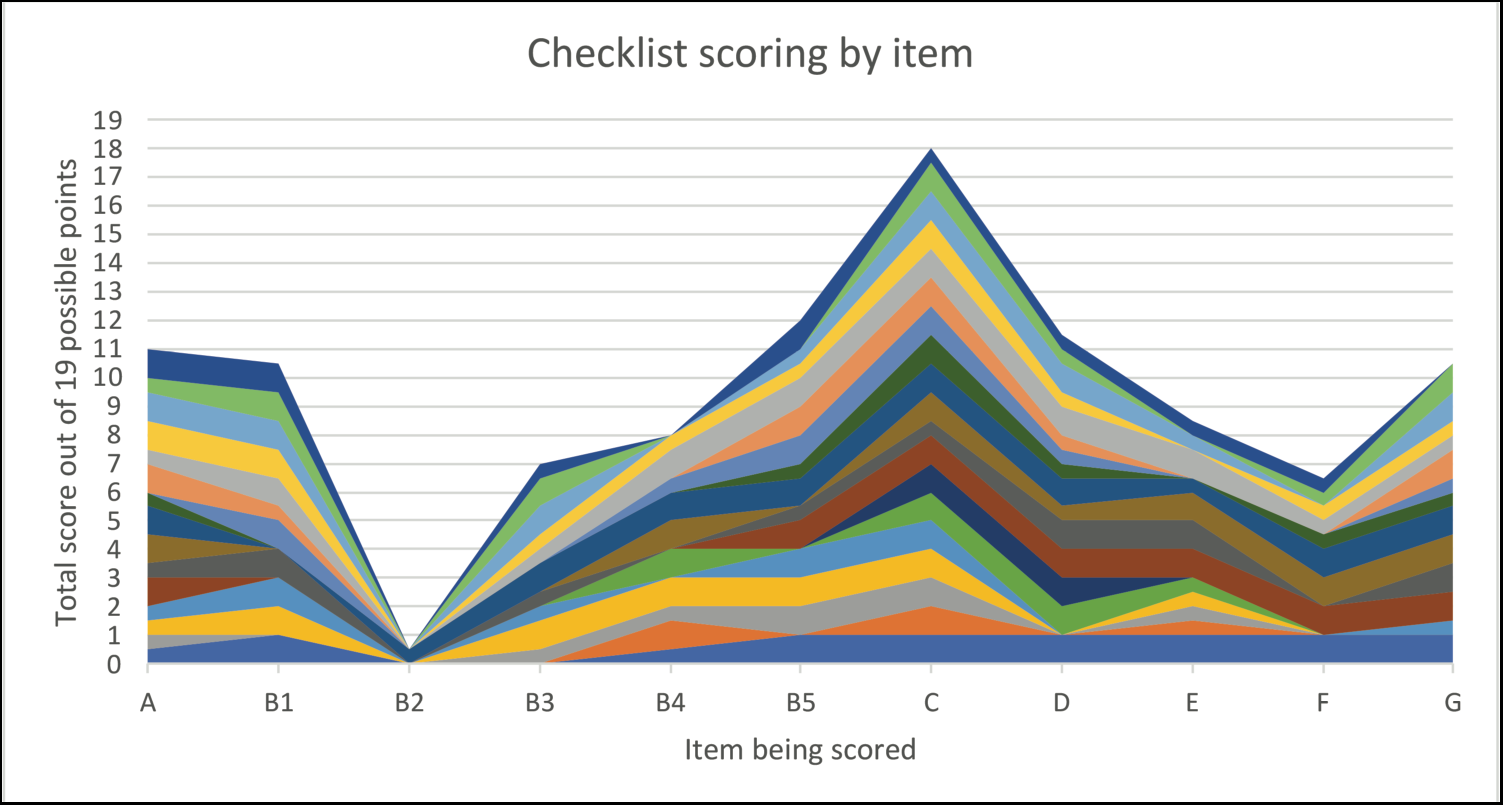
Number of studies reporting each checklist item. Each study is represented by a unique color on the mountain plot. Checklist items are labelled according to the original checklist (Supplemental Table 1).

Frequency and duration of the interventions were some of the most consistently reported details. Because of this, it was possible to discern that the frequency and duration of MBIs for AD/ADRD varied widely between the studies. No other details could be accurately compared because of the inconsistent item reporting and lack of detailed intervention descriptions across the studies. Specific songs or music genre used in the MBI was rarely reported, and the environment in which the music was delivered was rarely described. Within the manuscript text, the location of the specific details of the music interventions also varied. Most often, specific descriptions were reported in the methods section. However, some details were only found in the introduction or discussion sections, or could only be inferred from the descriptions as they were not overtly stated. This added difficulty when identifying whether a checklist item had been fulfilled, because it required careful and repeated reads through the papers to locate each specific item. Beyond the difficulty of locating the information, the specific details included in the intervention descriptions varied so greatly that precise replication of a reported intervention would be nearly impossible.

## DISCUSSION

There is favorable evidence that MBI’s produce beneficial outcomes for those living with AD/ADRD and their caregivers. However, the lack of consistency in which details are reported combined with the lack of detailed descriptions of the specific components of these MBI’s makes accurate reproduction of these interventions nearly impossible. Without the ability to accurately reproduce these interventions, validation of their results remains inconsistent. The specific music used in interventions was the most underreported checklist category, which is unfortunate because it is the foundation of a music-based intervention. Music is a diverse and general term, and even if a specific genre or song title is provided there is still variation in music components across performers and performances. These subtle musical variations could greatly influence the results of the intervention [25].

One of the most underreported items across studies was “B.2: Music”. Most studies described intervention duration, frequency, and group size, but few offered more than a vague description of music type, and specific songs and artists were rarely included. Several studies narrowed music intervention into active or passive categories, but these categories give little insight into the nature of the music itself. Most studies did not describe delivery volume, tempo, location or any other specifically descriptive categories. Replicating an MBI when the only details which have been provided are duration and frequency of the intervention is bound to result in a host of differing musical components which will cause changes in the effects produced. Providing specific references and descriptions of the music used in an intervention would allow more replicability for independent validation of an MBIs results.

Another highly underreported item, “A: Intervention Theory”, is also a crucial component of any study. Without a clear theory underlying the music intervention design, the biological mechanism being targeted, and the expected results; it is difficult to determine whether the intervention was truly effective. Studies frequently reported large conceptual domains that could be affected by music, such as memory or cognition, but rarely described how music specifically was hypothesized to affect a specific change. Results may be observed, but understanding what intervention component is producing them and what biological mechanism is being utilized is challenging if the theory has not been clearly defined and utilized in intervention design. This further contributes to the difficulty in replication and validation.

Limitations of this review include the search of only one database (PubMed) for records in English and one reviewer (BH) for record screening, as well as the limited number of AD/ADRD related MBI studies available for review. Rigor was increased by having two independent scorers (BH, AZ) and a third scorer (RL) to review and reconcile any interrater disagreement in scoring.

Our mapping of the current literature provides evidence that reporting music-based interventions with enough detail to replicate and validate the fidelity of interventions remains limited, thus restricting progress in the development and efficacy of music-based interventions for AD/ADRD. One goal of intervention research is to influence effective clinical practice. Translation of research into clinical practice has historically been lengthy, taking an average of 17 years [28]. Systematic reviews are one way to translate evidence-based research into clinical practice [29]; however, transparent reporting is needed within primary research studies to effectively support translation of clinical research to practice. According to Google Scholar, there are over 300 citations of the Reporting Guidelines for Music-based Interventions. The original article, published in 2011 in the *Journal of Health Psychology,* was also reprinted that same year in *Music and Medicine*, an interdisciplinary journal of the International Association of Music and Medicine that is specific to music-based intervention research and clinical practice [30]. The reporting guidelines for music-based interventions are easily accessible but have not been adopted as quickly as needed to align with trends in transparent reporting of health interventions, such as recommendations by the Equator Network [31]. We urge researchers to include music therapists or other music-based intervention experts into the conceptualization and operationalization of music-based interventions to ensure the intervention details are evident within the intervention manual and protocol implementation and to align with recommendations to advance rigor, replication and translation of music-based interventions [23, 25].

Replication and validation of results is a crucial component of scientific progress. A theory cannot be refined without repeated testing. A lack of clear and detailed descriptions of the theory behind an intervention design or the musical components of an intervention would be problematic if only one of these items was not reported. The lack of both of these items from a report makes replication and validation nearly impossible. The “Checklist for Reporting Music-Based Interventions” contains both of these items along with other important details and has been freely available since 2011. It was created with scientific rigor and for a specific purpose. Following this checklist will provide a framework to aid in consistent reproducibility of studies, and validation or invalidation of reported results. However, the checklist was not cited by any of the 19 studies we reviewed. If MBIs cannot be consistently validated their observed results will remain anecdotal in nature. To move MBIs from the realm of anecdotal evidence into the realm of prescriptive intervention, a consistent and ordered method of reporting is necessary. This method has already been provided, now is the time to put it to use.

## AUTHOR CONTRIBUTIONS

Rebecca Lepping (Conceptualization, Methodology, Validation, Formal analysis, Investigation, Resources, Writing – Original Draft, Writing – Review & Editing, Supervision, Funding acquisition); Benjamin Hess (Formal analysis, Investigation, Data curation, Writing – Original Draft, Writing – Review & Editing, Visualizaiton); Jasmine Taylor (Writing – Review & Editing); Deanna Hanson-Abromeit (Methodology, Writing – Review & Editing); Kristine Williams (Methodology, Writing – Review & Editing, Funding acquisition).

## ACKNOWLEDGEMENTS

The authors thank Ava Zatloukal (AZ) for assistance scoring the study articles.

## DATA AVAILABILITY

This review was not registered and a protocol was not prepared. The data supporting the findings of this study are available on request from the corresponding author.

## REFERENCES

[1] Groussard M, Chan TG, Coppalle R, Platel H (2019) Preservation of Musical Memory Throughout the Progression of Alzheimer’s Disease? Toward a Reconciliation of Theoretical, Clinical, and Neuroimaging Evidence. J Alzheimers Dis 68, 857–883.

[2] Gomez-Romero M, Jimenez-Palomares M, Rodriguez-Mansilla J, Flores-Nieto A, Garrido-Ardila EM, Gonzalez Lopez-Arza MV (2017) Benefits of music therapy on behaviour disorders in subjects diagnosed with dementia: a systematic review. Neurologia 32, 253–263.

[3] Soufineyestani M, Khan A, Sufineyestani M (2021) Impacts of Music Intervention on Dementia: A Review Using Meta-Narrative Method and Agenda for Future Research. Neurol Int 13, 1–17.

[4] Sarkamo T (2018) Cognitive, emotional, and neural benefits of musical leisure activities in aging and neurological rehabilitation: A critical review. Ann Phys Rehabil Med 61, 414–418.

[5] Dorris JL, Neely S, Terhorst L, VonVille HM, Rodakowski J (2021) Effects of music participation for mild cognitive impairment and dementia: A systematic review and meta-analysis. J Am Geriatr Soc 69, 2659–2667.

[6] Millan-Calenti JC, Lorenzo-Lopez L, Alonso-Bua B, de Labra C, Gonzalez-Abraldes I, Maseda A (2016) Optimal nonpharmacological management of agitation in Alzheimer’s disease: challenges and solutions. Clin Interv Aging 11, 175–184.

[7] Li K, Cui C, Zhang H, Jia L, Li R, Hu HY (2022) Exploration of combined physical activity and music for patients with Alzheimer’s disease: A systematic review. Front Aging Neurosci 14, 962475.

[8] Garcia-Navarro EB, Buzon-Perez A, Cabillas-Romero M (2022) Effect of Music Therapy as a Non-Pharmacological Measure Applied to Alzheimer’s Disease Patients: A Systematic Review. Nurs Rep 12, 775–790.

[9] Moreira SV, Justi F, Moreira M (2018) Can musical intervention improve memory in Alzheimer’s patients? Evidence from a systematic review. Dement Neuropsychol 12, 133–142.

[10] Popa LC, Manea MC, Velcea D, Salapa I, Manea M, Ciobanu AM (2021) Impact of Alzheimer’s Dementia on Caregivers and Quality Improvement through Art and Music Therapy. Healthcare (Basel) 9.

[11] Gomez-Gallego M, Gomez-Gallego JC, Gallego-Mellado M, Garcia-Garcia J (2021) Comparative Efficacy of Active Group Music Intervention versus Group Music Listening in Alzheimer’s Disease. Int J Environ Res Public Health 18.

[12] Moreno-Morales C, Calero R, Moreno-Morales P, Pintado C (2020) Music Therapy in the Treatment of Dementia: A Systematic Review and Meta-Analysis. Front Med (Lausanne) 7, 160.

[13] Lam HL, Li WTV, Laher I, Wong RY (2020) Effects of Music Therapy on Patients with Dementia-A Systematic Review. Geriatrics (Basel) 5.

[14] Ito E, Nouchi R, Dinet J, Cheng CH, Husebo BS (2022) The Effect of Music-Based Intervention on General Cognitive and Executive Functions, and Episodic Memory in People with Mild Cognitive Impairment and Dementia: A Systematic Review and Meta-Analysis of Recent Randomized Controlled Trials. Healthcare (Basel) 10.

[15] van der Steen JT, Smaling HJ, van der Wouden JC, Bruinsma MS, Scholten RJ, Vink AC (2018) Music-based therapeutic interventions for people with dementia. Cochrane Database Syst Rev 7, CD003477.

[16] Matziorinis AM, Koelsch S (2022) The promise of music therapy for Alzheimer’s disease: A review. Ann N Y Acad Sci 1516, 11–17.

[17] Fang R, Ye S, Huangfu J, Calimag DP (2017) Music therapy is a potential intervention for cognition of Alzheimer’s Disease: a mini-review. Transl Neurodegener 6, 2.

[18] Bleibel M, El Cheikh A, Sadier NS, Abou-Abbas L (2023) The effect of music therapy on cognitive functions in patients with Alzheimer’s disease: a systematic review of randomized controlled trials. Alzheimers Res Ther 15, 65.

[19] Lai X, Wen H, Li Y, Lu L, Tang C (2020) The Comparative Efficacy of Multiple Interventions for Mild Cognitive Impairment in Alzheimer’s Disease: A Bayesian Network Meta-Analysis. Front Aging Neurosci 12, 121.

[20] Lee S, Allison T, O’Neill D, Punch P, Helitzer E, Moss H (2022) Integrative review of singing and music interventions for family carers of people living with dementia. Health Promot Int 37, i49–i61.

[21] NIH NIoH, Notices of Funding Opportunities That Allow Music-Based Interventions, https://www.nccih.nih.gov/research/notices-of-funding-opportunities-that-allow-music-based-interventions,

[22] NIH NIoH, Dementia Care and Caregiver Support Intervention Research (R01 Clinical Trial Required), https://grants.nih.gov/grants/guide/pa-files/PAR-21-307.html,

[23] Edwards E, St Hillaire-Clarke C, Frankowski DW, Finkelstein R, Cheever T, Chen WG, Onken L, Poremba A, Riddle R, Schloesser D, Burgdorf CE, Wells N, Fleming R, Collins FS (2023) NIH Music-Based Intervention Toolkit: Music-Based Interventions for Brain Disorders of Aging. Neurology 100, 868–878.

[24] Hanson-Abromeit D (2015) A conceptual methodology to define the Therapeutic Function of Music. Music Therapy Perspectives 33, 25–38.

[25] Robb SL, Burns DS, Carpenter JS (2011) Reporting guidelines for music-based interventions. J Health Psychol 16, 342–352.

[26] Robb SL, Hanson-Abromeit D, May L, Hernandez-Ruiz E, Allison M, Beloat A, Daugherty S, Kurtz R, Ott A, Oyedele OO, Polasik S, Rager A, Rifkin J, Wolf E (2018) Reporting quality of music intervention research in healthcare: A systematic review. Complement Ther Med 38, 24–41.

[27] Page MJ, Moher D, Bossuyt PM, Boutron I, Hoffmann TC, Mulrow CD, Shamseer L, Tetzlaff JM, Akl EA, Brennan SE, Chou R, Glanville J, Grimshaw JM, Hrobjartsson A, Lalu MM, Li T, Loder EW, Mayo-Wilson E, McDonald S, McGuinness LA, Stewart LA, Thomas J, Tricco AC, Welch VA, Whiting P, McKenzie JE (2021) PRISMA 2020 explanation and elaboration: updated guidance and exemplars for reporting systematic reviews. BMJ 372, n160.

[28] Rubin R (2023) It Takes an Average of 17 Years for Evidence to Change Practice-the Burgeoning Field of Implementation Science Seeks to Speed Things Up. JAMA 329, 1333–1336.

[29] Tracy SL (2014) From bench-top to chair-side: How scientific evidence is incorporated into clinical practice. Dental Materials 30, 1–15.

[30] Robb SL, Burns DS, Carpenter JS (2011) Reporting Guidelines for Music-based Interventions. Music Med 3, 271–279.

[31] Network E, Enhancing the QUAlity and Transparency Of health Research, https://www.equator-network.org/reporting-guidelines/,

[32] Innes KE, Selfe TK, Brundage K, Montgomery C, Wen S, Kandati S, Bowles H, Khalsa DS, Huysmans Z (2018) Effects of Meditation and Music-Listening on Blood Biomarkers of Cellular Aging and Alzheimer’s Disease in Adults with Subjective Cognitive Decline: An Exploratory Randomized Clinical Trial. J Alzheimers Dis 66, 947–970.

[33] Innes KE, Selfe TK, Khalsa DS, Kandati S (2017) Meditation and Music Improve Memory and Cognitive Function in Adults with Subjective Cognitive Decline: A Pilot Randomized Controlled Trial. J Alzheimers Dis 56, 899–916.

[34] Innes KE, Selfe TK, Khalsa DS, Kandati S (2016) Effects of Meditation versus Music Listening on Perceived Stress, Mood, Sleep, and Quality of Life in Adults with Early Memory Loss: A Pilot Randomized Controlled Trial. J Alzheimers Dis 52, 1277–1298.

[35] Pongan E, Delphin-Combe F, Krolak-Salmon P, Leveque Y, Tillmann B, Bachelet R, Getenet JC, Auguste N, Trombert B, Dorey JM, Laurent B, Rouch I (2020) Immediate Benefit of Art on Pain and Well-Being in Community-Dwelling Patients with Mild Alzheimer’s. Am J Alzheimers Dis Other Demen 35, 1533317519859202.

[36] Kim D (2020) The Effects of a Recollection-Based Occupational Therapy Program of Alzheimer’s Disease: A Randomized Controlled Trial. Occup Ther Int 2020, 6305727.

[37] Park J, Tolea MI, Sherman D, Rosenfeld A, Arcay V, Lopes Y, Galvin JE (2020) Feasibility of Conducting Nonpharmacological Interventions to Manage Dementia Symptoms in Community-Dwelling Older Adults: A Cluster Randomized Controlled Trial. Am J Alzheimers Dis Other Demen 35, 1533317519872635.

[38] Baker FA, Lee YC, Sousa TV, Stretton-Smith PA, Tamplin J, Sveinsdottir V, Geretsegger M, Wake JD, Assmus J, Gold C (2022) Clinical effectiveness of music interventions for dementia and depression in elderly care (MIDDEL): Australian cohort of an international pragmatic cluster-randomised controlled trial. Lancet Healthy Longev 3, e153–e165.

[39] McCreedy EM, Sisti A, Gutman R, Dionne L, Rudolph JL, Baier R, Thomas KS, Olson MB, Zediker EE, Uth R, Shield RR, Mor V (2022) Pragmatic Trial of Personalized Music for Agitation and Antipsychotic Use in Nursing Home Residents With Dementia. J Am Med Dir Assoc 23, 1171–1177.

[40] Cheung DSK, Ho LYW, Chan LCK, Kwok RKH, Lai CKY (2022) A Home-Based Dyadic Music-with-Movement Intervention for People with Dementia and Caregivers: A Hybrid Type 2 Cluster-Randomized Effectiveness-Implementation Design. Clin Interv Aging 17, 1199–1216.

[41] Hsu MH, Flowerdew R, Parker M, Fachner J, Odell-Miller H (2015) Individual music therapy for managing neuropsychiatric symptoms for people with dementia and their carers: a cluster randomised controlled feasibility study. BMC Geriatr 15, 84.

[42] Feng L, Romero-Garcia R, Suckling J, Tan J, Larbi A, Cheah I, Wong G, Tsakok M, Lanskey B, Lim D, Li J, Yang J, Goh B, Teck TGC, Ho A, Wang X, Yu JT, Zhang C, Tan C, Chua M, Li J, Totman JJ, Wong C, Loh M, Foo R, Tan CH, Goh LG, Mahendran R, Kennedy BK, Kua EH (2020) Effects of choral singing versus health education on cognitive decline and aging: a randomized controlled trial. Aging (Albany NY) 12, 24798–24816.

[43] Mahendran R, Gandhi M, Moorakonda RB, Wong J, Kanchi MM, Fam J, Rawtaer I, Kumar AP, Feng L, Kua EH (2018) Art therapy is associated with sustained improvement in cognitive function in the elderly with mild neurocognitive disorder: findings from a pilot randomized controlled trial for art therapy and music reminiscence activity versus usual care. Trials 19, 615.

[44] Sarkamo T, Laitinen S, Numminen A, Kurki M, Johnson JK, Rantanen P (2016) Clinical and Demographic Factors Associated with the Cognitive and Emotional Efficacy of Regular Musical Activities in Dementia. J Alzheimers Dis 49, 767–781.

[45] Ray KD, Gotell E (2018) The Use of Music and Music Therapy in Ameliorating Depression Symptoms and Improving Well-Being in Nursing Home Residents With Dementia. Front Med (Lausanne) 5, 287.

[46] Liu MN, Liou YJ, Wang WC, Su KC, Yeh HL, Lau CI, Hu LY, Tsai SJ, Chen HY (2021) Group Music Intervention Using Percussion Instruments to Reduce Anxiety Among Elderly Male Veterans with Alzheimer Disease. Med Sci Monit 27, e928714.

[47] Pongan E, Tillmann B, Leveque Y, Trombert B, Getenet JC, Auguste N, Dauphinot V, El Haouari H, Navez M, Dorey JM, Krolak-Salmon P, Laurent B, Rouch I, Group LA (2017) Can Musical or Painting Interventions Improve Chronic Pain, Mood, Quality of Life, and Cognition in Patients with Mild Alzheimer’s Disease? Evidence from a Randomized Controlled Trial. J Alzheimers Dis 60, 663–677.

48. [48] de la Rubia Orti JE, Garcia-Pardo MP, Iranzo CC, Madrigal JJC, Castillo SS, Rochina MJ, Gasco VJP (2018) Does Music Therapy Improve Anxiety and Depression in Alzheimer’s Patients? J Altern Complement Med 24, 33–36.

[49] Kwak J, Anderson K, O’Connell Valuch K (2020) Findings From a Prospective Randomized Controlled Trial of an Individualized Music Listening Program for Persons With Dementia. J Appl Gerontol 39, 567–575.

